# Clinical characteristics of Chilean patients with rheumatic diseases and COVID-19: data from the Covid-19 Global Rheumatology Alliance physician-reported registry

**DOI:** 10.1101/2021.12.07.21267429

**Authors:** Oriela Martínez, Francisca Valenzuela, Sebastián Ibáñez

## Abstract

**Objective:** The coronavirus disease 2019 (COVID-19) pandemic, caused by the severe acute respiratory syndrome coronavirus 2 (SARS-Cov-2), has registered more than 234 million confirmed cases and more than 4.7 million deaths throughout the world until October 2, 2021. During the last few months, a significant number of reports of COVID-19 in patients with rheumatic diseases have been published. In this study the objective is to report the clinical characteristics of Chilean patients with rheumatic diseases and COVID-19 reported in the “Global Rheumatology Alliance” (GRA) physician registration platform.

**Methods:** Chilean patients with rheumatic diseases and COVID-19 were included in the Covid-19 GRA physician-reported registry.

**Results:** 54 patients were included. The most common primary rheumatic disease was rheumatoid arthritis (RA) with 28 cases (51.9%). 30 patients (55.6%) used corticosteroids, of which 20 (66.7%) used a dose of 10 mg or less. 33 patients (61.1%) only used conventional DMARDs, 4 (7.4%) only biological, and 6 (11.1%) the combination. A total of 35 patients (64.8%) had to be hospitalized. 2 patients (3.7%) died. 26 patients of the 35 hospitalized (74.2%) required some type of ventilatory support, of which 5 (19.2%) required non-invasive and 8 (30.8%) invasive mechanical ventilation or extracorporeal membrane oxygenation (ECMO).

**Discussion:** Most of included Chilean rheumatic patients were hospitalized, with a low mortality rate but with a high percentage of patients requiring at least non-invasive mechanical ventilation.

**Key Points:**

- The most common primary rheumatic disease was rheumatoid arthritis (RA) followed by lupus (LES)
- Most of the included Chilean rheumatic patients were hospitalized, with a high percentage of patients requiring at least non-invasive mechanical ventilation, but with a low mortality rate.
- Worsening of arthralgias or activation of the rheumatic disease was not reported.

## Introduction

The coronavirus disease 2019 (COVID-19) pandemic, caused by the severe acute respiratory syndrome coronavirus 2 (SARS-Cov-2), has registered more than 234 million confirmed cases and more than 4.7 million deaths throughout the world until October 2, 2021, according to data published by the Johns Hopkins University (1). In Latin America, according to the World Health Organization, by October 2, 2021, almost 47 million cases had been registered, with approximately 1.5 million deaths (2), while Chile reported a total of 1.6 million cases with 37 thousand deaths from COVID-19 (3).

COVID-19 can generate a severe and aggressive illness, requiring hospitalization and ventilatory support, and potentially resulting in death. During the last few months, a significant number of case series and reports of COVID-19 in patients with rheumatic diseases have been published; one of the most important being the registry of the “COVID-19 Global Rheumatology Alliance” (GRA), the response of the rheumatology community to the global pandemic of COVID-19 (4). Since its launch in March 2020, more than 19,000 cases have been reported worldwide through its European and World-wide registries. In this study the objective is to report the clinical characteristics of Chilean patients with rheumatic diseases and COVID-19 reported in the GRA physician registration platform.

## Methods

Chilean patients with rheumatic disease and COVID-19 were included in the physician registry of the GRA between April 2020 and August 2021. COVID-19 diagnostic method as well as sociodemographic information, including age, sex, smoking status, rheumatic diagnosis, disease activity, comorbidities, and pre-COVID-19 medications were recorded. Medications prior to COVID-19 were classified as conventional synthetic disease-modifying antirheumatic drugs (DMARDs), biological DMARDs, synthetic plus biological DMARDs, and use of corticosteroids. Regarding COVID-19, the duration of symptoms, the status of the infection (resolved or not resolved at the time of the report, or unknown), the use of a specific drug treatment for COVID-19, the need for ventilatory support, infection outcome [death, hospitalization, complications such as acute respiratory distress syndrome (ARDS), sepsis, myocarditis / heart failure, secondary infection, cytokine storm] were also recorded.

### Statistical analysis

The mean and standard deviation (SD) were calculated for the variables with normal distribution, and the median and interquartile range (IQR) were calculated for the variables without normal distribution. All analyzes were performed using the IBM SPSS Statistics version 24 program for Macintosh.

### Ethical Considerations

This study was approved by the Department of Academic Development and Research and the Scientific Ethics Committee of the “Clinica Alemana-Universidad del Desarrollo” Medicine Faculty.

## Results

54 patients were included, from April 2020 to August 2021. Of the total, 41 patients (75.9%) were from the metropolitan region. The mean age of the patients was 53.4 years (SD 14.6), and 36 patients (66.7%) were women. The most common primary rheumatic disease was rheumatoid arthritis (RA) with 28 cases (51.9%), followed by lupus (LES) with 7 cases (13%). At the time of COVID-19 diagnosis, 30 patients (55.6%) were in remission or had minimal or low activity of their underlying rheumatic disease, 8 patients (14.8%) had moderate activity and 8 (14.8%) severe activity, according to the physician appreciation. 30 patients (55.6%) used corticosteroids, of which 20 (66.7%) used a dose of 10 mg or less, and in 18 (60%) the dose was increased after COVID-19 diagnosis. 33 patients (61.1%) used only conventional DMARDs, 4 (7.4%) only biologics, and 6 (11.1%) the combination. DMARDs were stopped in 22 patients (51.2%) after COVID-19 diagnosis. Regarding comorbidities, 28 patients (51.9%) were hypertensive, 9 (16.7%) had underlying lung disease, 7 (13%) had diabetes, 1 (1.9%) had a body mass index (BMI) greater than 40. 13 patients (24.1%) had ever smoked.

Regarding COVID-19 place of diagnosis, 22 cases (40.7%) were diagnosed in the emergency service, 20 (37%) in the outpatient setting and 10 (18.5%) during hospitalization. 31 cases (57.4%) were infected by being close contact of a confirmed or probable case, and 12 (22.2%) reported as a possible place of contagion attending a health care center where COVID-19 cases were handled. In 47 (87%) patients the diagnosis was confirmed by polymerase chain reaction (PCR), 4 cases (7.4%) were diagnosed only by symptoms and 3 cases (5.6%) by a compatible CT scan. 52 cases (96.3%) were symptomatic, the main symptoms being fever in 29 patients (53.7%), cough also in 29 (53.7%), and dyspnea in 27 cases (50%) (table 1).

**Table 1.** Demographic characteristics of patients with rheumatological diseases and COVID 19 (n = 54)

|  |  |
| --- | --- |
| ***Women, n(%)*** | **36 (66.7)** |
| ***Age (SD)*** | **53.4 (14.6)** |
| ***Primary rheumatologic diagnosis, n (%)*** |  |
| **RA** | **28 (51.9)** |
| **SLE** | **7 (13)** |
| **Spondyloarthritis** | **4 (7.4)** |
| **Sjogren's syndrome** | **3 (5.6)** |
| **Inflammatory myopathies** | **3 (5.6)** |
| **Other** | **7 (12.9)** |
| ***Comorbidities, n (%)*** |  |
| **HT** | **28 (51.9)** |
| **Lung disease** | **9 (16.7)** |
| **Diabetes** | **7 (13)** |
| **Cancer** | **3 (5.6)** |
| **CVA** | **3 (5.6)** |
| ***Ever smokers, n (%)*** | **13 (24.1)** |
| ***COVID-19 diagnostic method, n (%)*** |  |
| **PCR** | **47 (87)** |
| **Based on symptoms** | **4 (7.4)** |
| **CT scan** | **3 (5.6)** |
| ***Symptoms, n (%)*** |  |
| **Fever** | **29 (53.7)** |
| **Cough** | 29 (53.7) |
| **Dyspnoea** | 27 (50) |
| **Myalgia** | 25 (46.3) |
| **Headache** | 22 (40.7) |
| **Diarrhea, nausea, vomiting** | 16 (29.6) |
| **Odynophagia** | 14 (25.9) |
| **Anosmia** | 13 (24.1) |
| **Dysgeusia** | 12 (22.2) |
| **Arthralgia** | 2 (3.7) |
| *Received specific treatment for COVID-19,*<br>*n (%)* | 30 (55.6) |
| **Corticosteroids** | 16 (53.3) |
| **Azithromycin** | 8 (26.7) |
| **Antimalarials** | 6 (20) |

SD: standard deviation RA: rheumatoid arthritis SLE: Systemic lupus erythematosus HT: arterial hypertension CVA: cerebrovascular accident BMI: body mass index PCR: polymerase chain reaction CT scan: computerized axial tomography DMARD: conventional synthetic disease-modifying antirheumatic drugs.
The "other" rheumatic disease category included polymyalgia rheumatica, vasculitis, psoriatic arthritis, antiphospholipid syndrome, other inflammatory arthritis, undifferentiated connective tissue disease

**Table 2.**
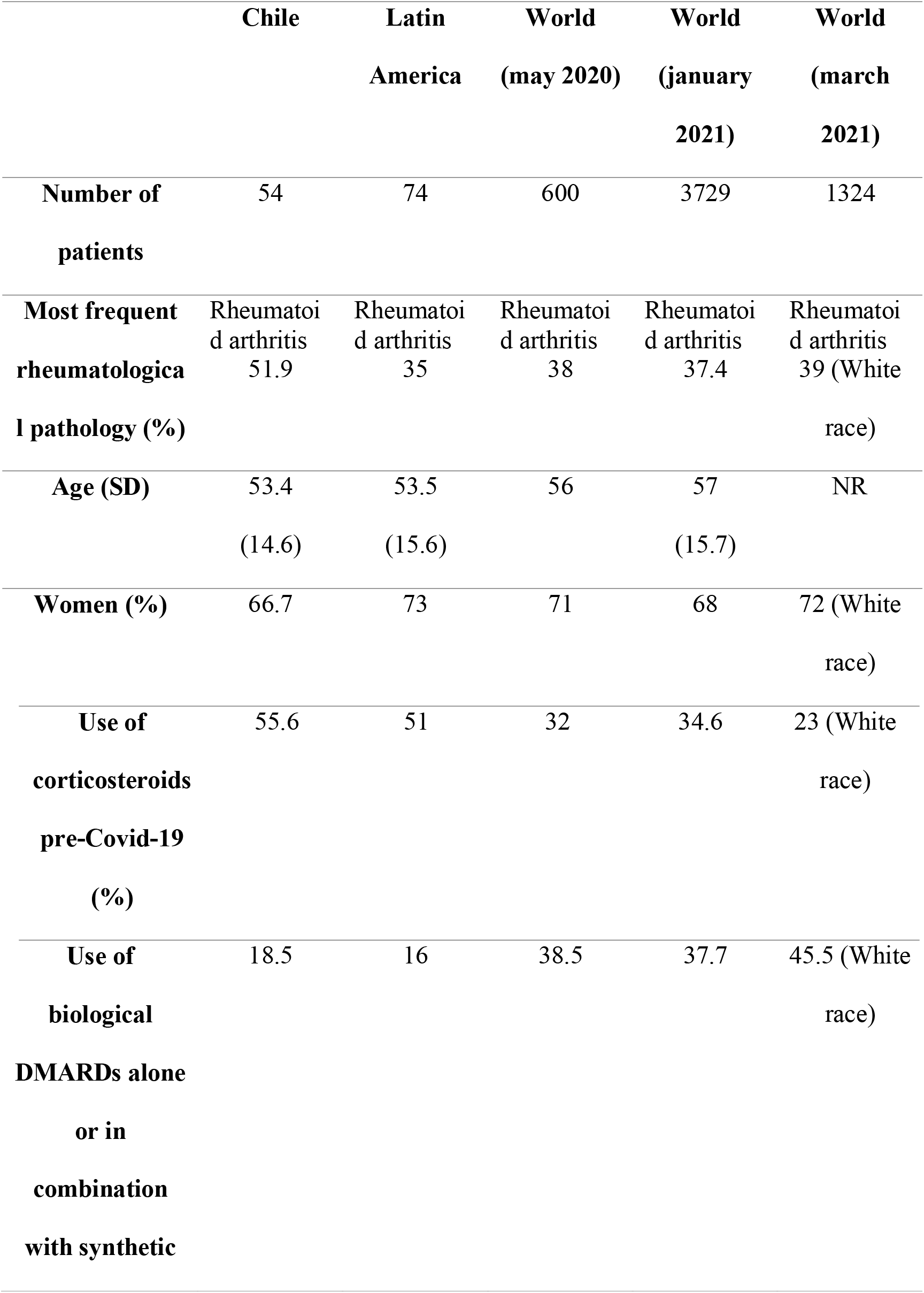
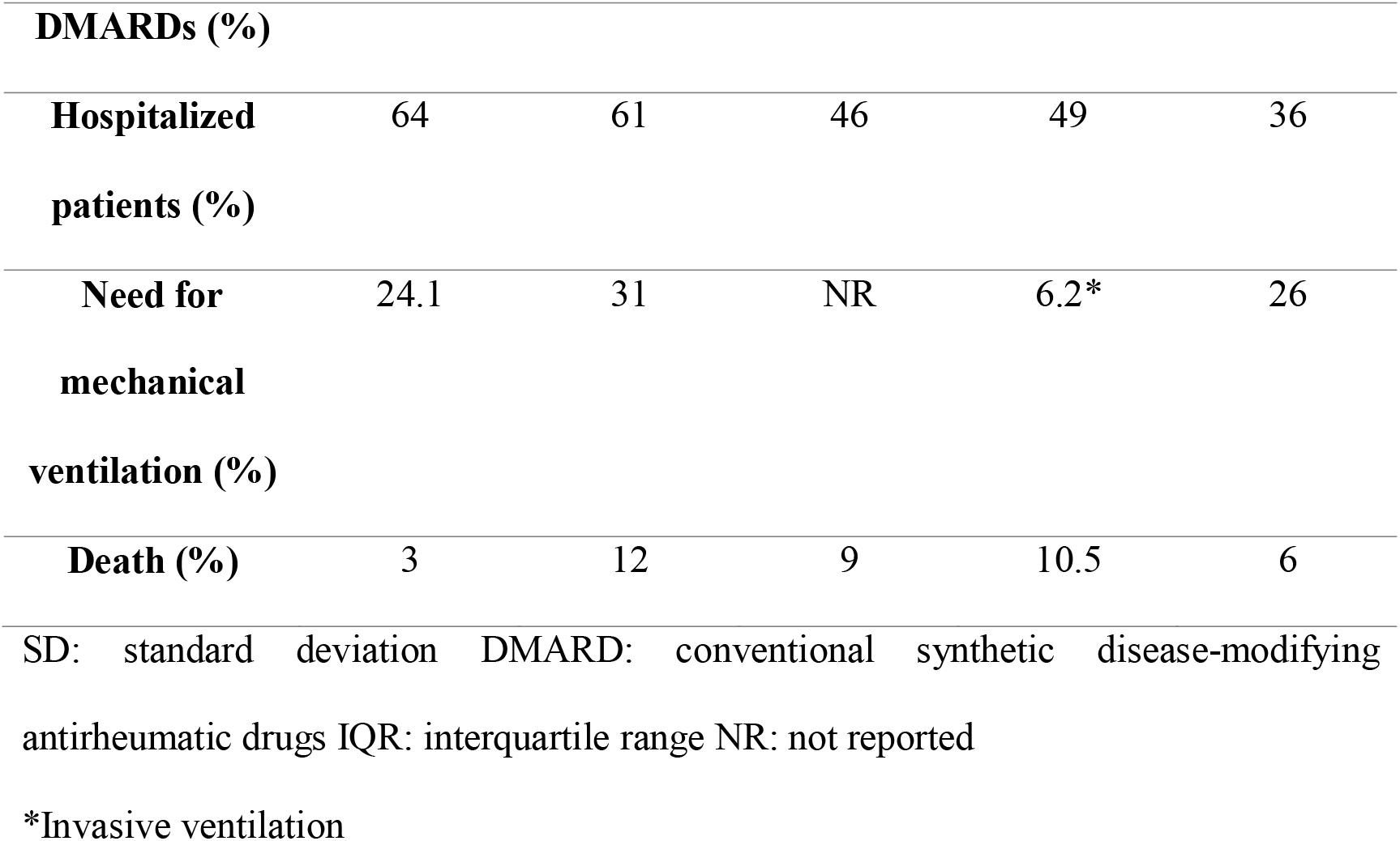
Characteristics of the Chilean, Latin American and world patients of the Global Alliance of Rheumatology COVID-19.

Specific drug treatment for COVID-19 was used in 30 patients (55.6%), of these, 16 (29.6%) received corticosteroids, 8 (14.8%) received azithromycin, 6 (11.1%) antimalarials, and 1 (1.9%) received tocilizumab. In 18 of the 30 patients (60%) who used corticosteroids before the diagnosis of COVID-19, the dose was increased after the diagnosis. At the time of registering, 40 patients (76.9%) had resolved symptoms. The duration of symptoms in these patients was a median of 21 days (interquartile range (IQR) 10-31.5). A total of 35 patients (64.8%) had to be hospitalized. 2 patients (3.7%) died. Among the 35 hospitalized patients, 26 (74.2%) required some type of ventilatory support, 13 (24.1%) at least non-invasive mechanical ventilation and of these 8 (14.8%) patients required invasive mechanical ventilation (IMV) or extracorporeal membrane oxygenation (ECMO). None of the registered patients presented a cytokine storm like syndrome.

## Discussion

Since the COVID-19 pandemic began, the question has arisen as to whether patients with pre-existing inflammatory diseases mediated by the immune system, such as rheumatic diseases, are at increased risk of SARS CoV2 infection or severe COVID-19 outcomes. The first GRA report was published in may 2020 (5), where the factors associated with hospitalization were examined in 600 rheumatic patients with COVID-19 from 40 countries, of which 46% (277 patients) required hospitalization. Both, advanced age and the presence of previous comorbidities (arterial hypertension, previous lung disease, diabetes mellitus), were factors associated with hospitalization (as in the general population). The most frequent underlying rheumatic disease was rheumatoid arthritis followed by systemic lupus erythematosus. The use of high-dose glucocorticoids (≥10 mg per day of prednisone equivalent) was associated with an increased risk of hospitalization. The study also included a preliminary analysis of biological DMARD exposure and found that compared to patients who were not receiving biological DMARDs, they were less likely to be hospitalized, regardless of the presence of other risk factors. In the Chilean registry, the inclusion of hospitalized patients was favored, and the number of registered cases was low, so performing analysis of associations of risk for hospitalization and worse outcome of infection did not seem appropriate to us. The median of days from onset to resolution of symptoms was 13 days in the global report, while in Chilean patients it was 21 days.

In September 2020 another GRA study was published comparing the characteristics of 74 patients with rheumatic diseases and COVID-19 reported in Latin America with 583 from the rest of the world. The most frequent rheumatic diseases in both publications were rheumatoid arthritis and systemic lupus erythematosus, and mortality was similar in both groups (12% in Latin America vs 11% in the rest of the world). However, the Latin American registry included a higher percentage of hospitalized patients (61% in Latin America vs 45% in the rest of the world) and a high percentage needed at least non-invasive ventilation. This was similar to our Chilean report, but the reported mortality was higher in the Latin American cohort.

In January 2021, a publication derived from the GRA (7) registry determined the factors associated with death related to COVID-19 in people with rheumatic disease. Of 3729 patients, 1739 (49%) was hospitalized, 187 (6.2%) required invasive ventilation and 390 (10.5%) died. Death was associated with known general factors (older age, male sex and specific comorbidities) and disease-specific factors (disease activity and specific medications). The association with moderate/high disease activity highlights. Caution may be required with rituximab, sulfasalazine, glucocorticoid dosages and some immunosuppressants. Mortality was higher, but the percentage of hospitalized patients was lower in this study than that reported in the Chilean registry (10.5% vs 3% deaths, 49% vs 64% hospitalized patients).

A more recent publication from the GRA (8), with a total of 1,324 patients, concluded that African American, Latinos, and Asian patients were more likely to require hospitalization compared to white patients. Latino patients were also 3 times more likely to require ventilatory support. We think these differences may be due in large part to a bias in the registry, clearly in Chile the registry of hospitalized patients was favored. No differences in mortality were found based on race / ethnicity. The results of this study are similar to the Chilean data in terms of mortality and the use of at least non-invasive mechanical ventilation reported, despite the fact that in Chile the registration of hospitalized patients was favored (64% Chile vs 36% Global).

This study had some limitations, such as the difficulty in collecting data, due to medical exhaustion, the increase in the care burden in health centers, constant changes in the treatment paradigm, therefore, the number of patients included is low compared to those affected by COVID-19 in Chile. Therefore, we did not carry out other association analyzes, as they would not reflect the reality of what happened. It is evident that the median of symptoms is higher than that reported in other studies, this may be because the patients with more days of symptoms were those who attended health centers and were hospitalized. In Chilean rheumatic patients, COVID-19 symptoms were the classic symptoms described for the disease, a low percentage manifested arthralgia, and activation of the rheumatic disease was not reported. Less use of corticosteroids and greater use of biological DMARDS are evidenced in rheumatic patients from the rest of the world compared to Chilean patients, possibly related to accessibility given by economic factors and health policies of each region. Regarding the specific treatment for COVID 19, some patients were treated with therapies that were initially believed to work, but later evidence showed that they did not, such as hydroxychloroquine or azithromycin. Only one patient received tocilizumab, probably because of the time of registration (early in the pandemic). It is not mentioned whether the patients were vaccinated because when the data collection began, it was not yet available, and Chile began the vaccination process in patients with comorbidities in February 2021, and very few patients were registered after that.

It is concluded that most of the Chilean rheumatic patients included were hospitalized with a low mortality rate, although with a high percentage of patients requiring at least non-invasive mechanical ventilation

## Data Availability

All data produced in the present study are available upon reasonable request to the authors

